# The genetic architecture of medication-use

**DOI:** 10.1101/2020.09.30.20204438

**Authors:** Palle Duun Rohde, Peter Sørensen, Mette Nyegaard

## Abstract

Genomics has been forecasted to revolutionise human health by improving medical treatment through a better understanding of the molecular mechanisms of human diseases. Despite great successes of the last decade’s genome-wide association studies (GWAS), the results have to a limited extent been translated to genomic medicine. We propose, that one route to get closer to improved medical treatment is by understanding the genetics of medication-use. Here we obtained entire medication profiles from 335,744 individuals from the UK Biobank and performed a GWAS to identify which common genetic variants are major drivers of medication-use. We analysed 9 million imputed genetic variants, estimated SNP heritability, partitioned the genomic variance across functional categories, and constructed genetic scores for medication-use. In total, 59 independent loci were identified for medication-use and approximately 18% of the total variation was attributable to common genetic (minor allele frequency >0.01) variants. The largest fraction of variance was captured by variants with low to medium minor allele frequency. In particular coding and conserved regions, as well as transcription start sites, displayed significantly enrichment of heritability. The average correlation between medication-use and predicted genetic scores was 0.14. These results demonstrate that medication-use *per se* is a highly polygenic complex trait and that individuals with higher genetic liability are on average more diseased and have a higher risk for adverse drug reactions. These results provide an insight into the genetic architecture of medication use and pave the way for developments of multicomponent genetic risk models that includes the genetically informed medication-use.

## Introduction

Understanding the relationship between DNA sequence variation and the predisposition to common diseases has interested researchers for decades. In particular after the initial release of the human genome (1), the number of polymorphic genetic variants associated with disease predisposition has grown exponentially to more than 60,000 associations (2–4). Genome-wide association studies (GWAS) have provided new insight into the biology and genetic epidemiology of many human complex diseases, which is essential for innovative developments within genomic medicine.

The traditional *one-size-fits-all* approach for disease diagnosis and treatment has been proven to be inefficient, expensive and sometimes with adverse clinical reactions. Personalised medicine or genomic medicine, which often are used interchangeably (5), is foreseen to change the way we prevent, diagnose and treat medical conditions (6,7). The goal of genomic medicine is to develop targeted preventive or treatment approaches based on the individual’s genetic makeup, environmental exposures and lifestyle parameters (6,7). Fundamental for development of genomic medicine is accurate knowledge about the disease pathogenesis. Equally important is the realisation of the genetic contributions to drug response *per se* (8–10). Genetic variation among patients modulates the drug efficiency and can impose toxic effects (adverse drug reactions) (11,12). Therefore, understanding the genetic architecture of drug response is absolutely essential for the development of genomic medicine.

A major challenge and hindrance in studying genetic factors influencing drug response variability is the lack of accessible data. Despite the emerge of large biobanks such as the United Kingdom Biobank (13), Japan Biobank (14) and Estonia Biobank (15), that contain genetic and deep phenotypic information of the participants, information on response to medical treatment is absent. The accessibility of electronic health records and self-reported health status may provide means for alternative approaches for studying the genetic basis of traits of relevance for medication-use.

Previously, Wu *et al*. (2019)(16) performed a genetic analysis of medication-use in the United Kingdom Biobank (UKBB). They categorised medications based on the drugs active substances according to the organ or system they act on and their pharmacological properties. They performed a genetic analysis on 23 isolated groups of medications and identified a very large number of independent loci associated with the different drug categories. Instead of grouping drugs based on the active substances and analysing those individually, we investigated the entire medication profile of individuals from the UKBB. The aim of the current study was to investigate the genetic basis of self-reported medication-use in the UKBB. We defined medication-use as the total number of different prescription and over-the-counter medications individuals were taking at the time of the verbal interview with the UKBB assessment centres. Medication-use – as defined above – has the clear advantage that it is easily quantifiable compared to for example drug efficiency. Assuming current medication-use as a quantitative trait phenotype we conducted a genetic analysis of 335,744 unrelated individuals from the UKBB. We hypothesised that across medical conditions, medication-use has a detectable genetic component, and expected medication-use to be genetically correlated with common diseases as commonly prescribed drugs is likely to be a proxy for major disease groups. Extensive medication-use has in individuals older than 65 years been shown to be associated with ill health and morbidity (17,18); hence, we further hypothesised that medication-use was genetically correlated with health-related outcomes. If medication-use has a genetic component, understanding the genetic basis is important because many medications has side effects, and increased drug usage might be associated with higher risk of toxic effects. Hence, genetic predisposition to high medication-use could be used as guidance in treatment plans aiming to reduce the total number of medications.

## Results

We performed a genetic analysis of current medication-use within the white British cohort of the UK Biobank (*n*=335,744). We defined current medication-use as the number of different prescription and over-the-counter medicines the participants were taking regularly at the time of the verbal interview (short term medication, like antibiotics or analgesics was not included). The average number of medications taken by males were 2.34 (standard deviation (sd) 2.7) and 2.67 (sd 2.7) for females, with a linear increase in the number of medications taken with increasing age (Supplementary Figure S1). Interestingly, the mean number of drugs taken for individuals with an ICD10-code for adverse drug reaction (Supplementary Table S1)(19) was significant larger (mean=4.09, sd=3.57) than individuals without such diagnoses (mean=2.23, sd=2.43; t-test=-101.77, df=46662, *P*<2.2×10^−16^), suggesting that individuals taking a larger number of mediations are more likely to encounter an adverse drug reaction or that individuals experiencing adverse drug reactions are harder to treat, requiring more medication. The participants have reported a total of 3,247 different medications, where the most frequently used drugs were paracetamol (n=61,604), aspirin (n=44,894), ibuprofen (n=41,756) and simvastatin (n=38,379) (Supplementary Table S2).

After SNP quality control (see Methods) there were 9,804,629 autosomal SNPs left for GWAS analysis. In total, we identified 59 independent quantitative trait loci for current medication-use (Fig. 1, Supplementary Table S3). The strongest associated locus was within the human leucocyte antigens (HLA) complex (rs35248896, *P*=1.52×10^−46^). Because of the complexity of the HLA region (20,21) we only allowed one significant locus at this genomic region (additional three loci passed the significance threshold within the HLA region but were excluded, Supplementary Table S3). Among the 59 genome-wide significant loci 14 of them were located in intergenic regions.

**Figure 1.**
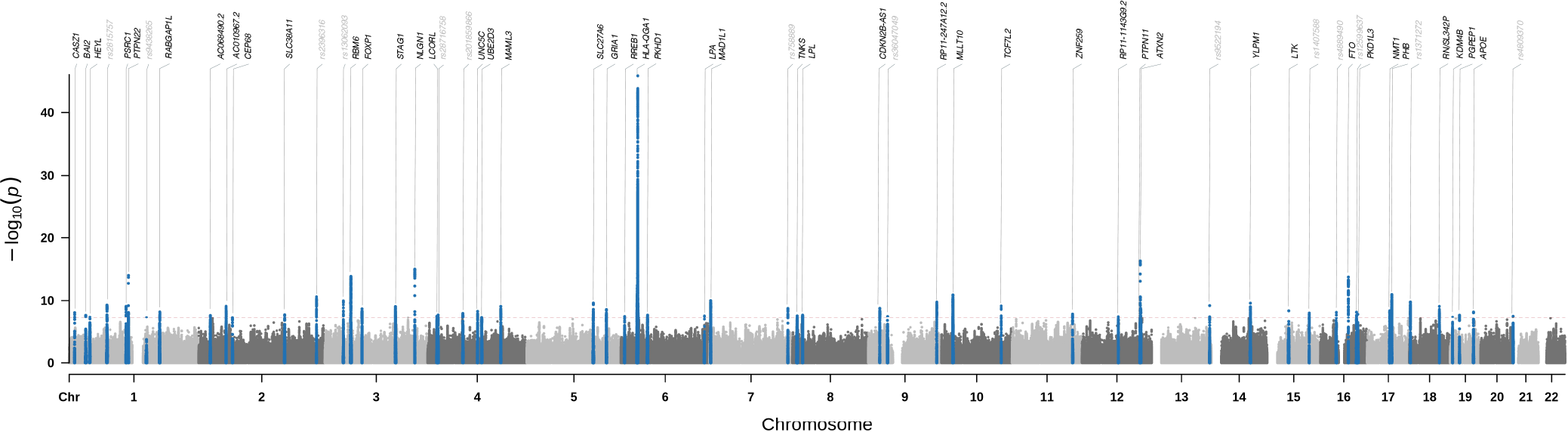
Manhattan plot for medication-use in the UKBB (n=335,744). The x-axis is chromosomal position, and the y-axis shows the negative logarithm base-10 to the *P* values from regression of current medication-use on 9,804,629 SNPs. The horizontal red line shows the genome-wide significance level (5×10^−8^). Independent genome-wide significant loci (within 1000 kb and r^2^<0.01) are depicted in blue. For each significant locus the gene within 2000kb is shown (for intergenic loci the lead SNP is shown).

Using the medication-use GWAS summary statistics (excluding the HLA region) we estimated the proportion of variation in medication-use explained by the SNPs 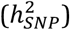 to 0.18±0.005. Next, we partitioned the total heritability to heritability captured by individual autosomal chromosomes and found a linear association between the proportion of heritability captured by each autosomal chromosome and the number of SNPs per chromosome (R^2^=0.9, Fig. 2A) suggesting that medication-use is a highly polygenic trait. The genomic variance explained per variant within minor allele frequency bins indicate that low frequent variants capture about 3 times more genetic variance than higher frequent variants does (Fig. 2B). Also, we estimated the enrichment score across 24 functional categories (obtained from Finucane et al (2015) (22)), which is the estimated share of 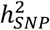 divided by its expected share under the assumed heritability model (22,23) (Fig 2C). In particular, the conserved genomic region and transcription start sites (TSS) were highly enriched accounting for 4.5% and 1.3% of 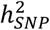, respectively.

**Figure 2.**
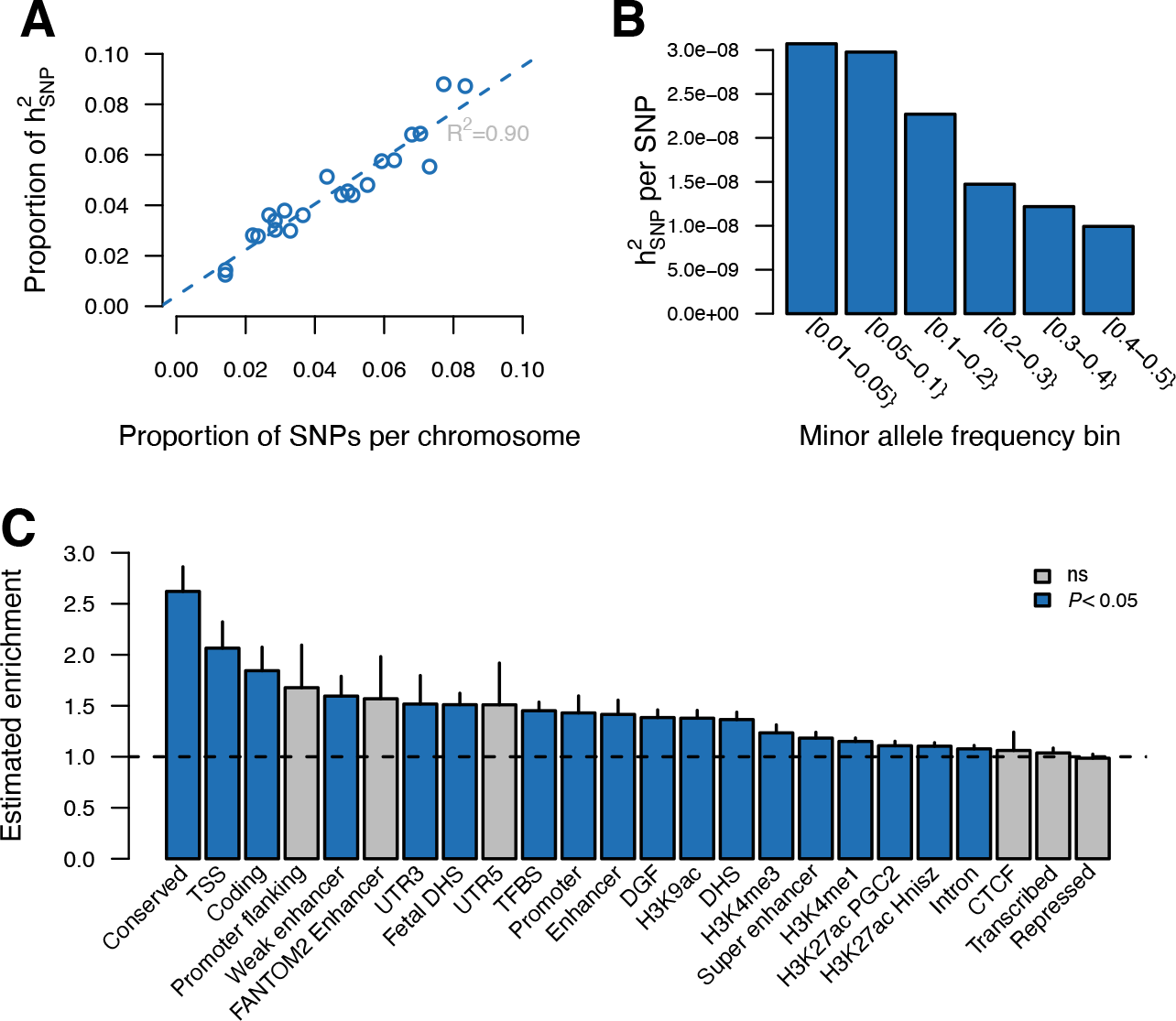
Partitioning of genomic variance for medication-use. **A:** Proportion of genomic variance captured per autosomal chromosome as function of the proportion of SNPs per chromosome. **B**: Proportion genomic variance, scaled by the number of SNPs, captured by minor allele frequency. **C:** Estimated enrichment score for functional categories. Vertical line segments mark the standard deviation on enrichment score. Horizontal dashed line marks an enrichment score of no enrichment. CTCF: a highly conserved multifunctional DNA-binding protein, DGF: digital genomic footprint, DHS: DNase I hypersensitivity sites, TFBS: transcription factor binding site, TSS: transcription start site.

We constructed genetic scores for medication-use by re-estimating the SNP-effects using a five-fold cross validation scheme. The scoring was performed on five levels of LD pruning (r^2^) and across eleven *P*-value thresholds. The maximum prediction accuracy (correlation between genetic scores and medication-use ∼0.14) was obtained when removing markers with r^2^>0.5 (Supplementary Fig. S2) at a *P*-value of 0.9 (Fig. 3A), which includes about 1.5 million genetic markers (Supplementary Fig. S3). Stratifying individuals based on their genetic score we see that individuals within the top 5% highest genetic score has increased medication-use compared with individuals with the 5% lowest genetic score (Fig. 3B, Supplementary Fig. S4). Moreover, individuals with the 5% highest genetic scores has significant more ICD10 diagnoses than those individuals with the 5% lowest genetic scores (10.8 diagnoses and 6.4 diagnoses, respectively, Supplementary Fig. S5). There is, however, no visual difference in which diseases they have been diagnosed with (Supplementary Fig. S6); individuals with the highest genetic score for medication-use simply has more diagnoses than those with low genetic scores (Supplementary Fig. S5B and Fig. S6). Using the classification of adverse drug reactions from Hohl *et al*. (2014) (19) (Supplementary Table S1), we further see, that individuals with predicted high risk and an experience of adverse drug reactions on average have a 1.6 fold higher medication-use compared with low risk individuals (Supplementary Fig. S7 and Table S5).

**Figure 3.**
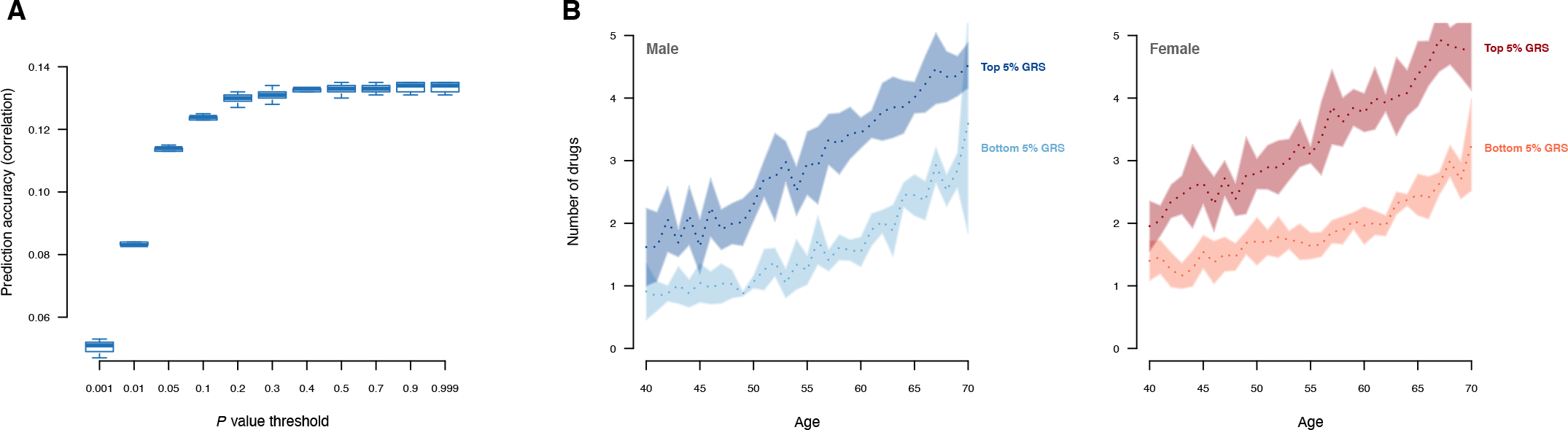
Prediction of number of medications used. **A:** Prediction accuracy (measured by the correlation between observed and predicted values) for medication-use across the range of *P* values. Results shown is for r^2^<0.5 as it gave the highest accuracy across different r^2^-values (see Supplementary Figure S2 for all r^2^ values). **B:** Averaged number of drugs (over the five training sets) used by males and females stratified by top 5% and bottom 5% of genetic scores. Shadings corresponds to the standard error over the five training sets.

Finally, we computed the genetic correlation between medication-use and 257 quantitative traits and complex diseases using LD Hub (24). We found a significant genetic correlation with 115 traits (Bonferroni adjusted *P* value <0.01) across 26 categories, except bone traits, where no genetic correlation with medication use was found (Fig. 4, Supplementary Table S4). Expectedly, medication-use was positively genetically correlated with major common complex diseases in particularly coronary artery disease, type 2 diabetes, asthma, lung cancer and major depressive disorder (Fig. 5). Parents age at death was the trait that was most negatively genetically correlated with medication-use indicating that higher medication use correlate with lower age at death (higher mortality) of parents (Fig. 5). We also found that the number of years in school and if completed college were negatively correlated with medication-use. Number of cigarettes smoked per day and medication-use was positively genetically correlated, and medication-use was also genetically correlated with sleep traits; insomnia and sleep duration (Fig. 5). Finally, we estimated genetic correlations between medication-use and the medication categories previously published by Wu *et al*. (2019) (16) (Supplementary Figure S8). The average genetic correlation between our definition of medication-use and the 23 medication categories was 0.52 (sd 0.22; Supplementary Table S6), and the category ‘drugs affecting bone structure and mineralization’ was the only insignificant result, which is in agreement with the observation that medication-use was not genetically correlated with any bone traits (Fig. 4).

**Figure 4.**
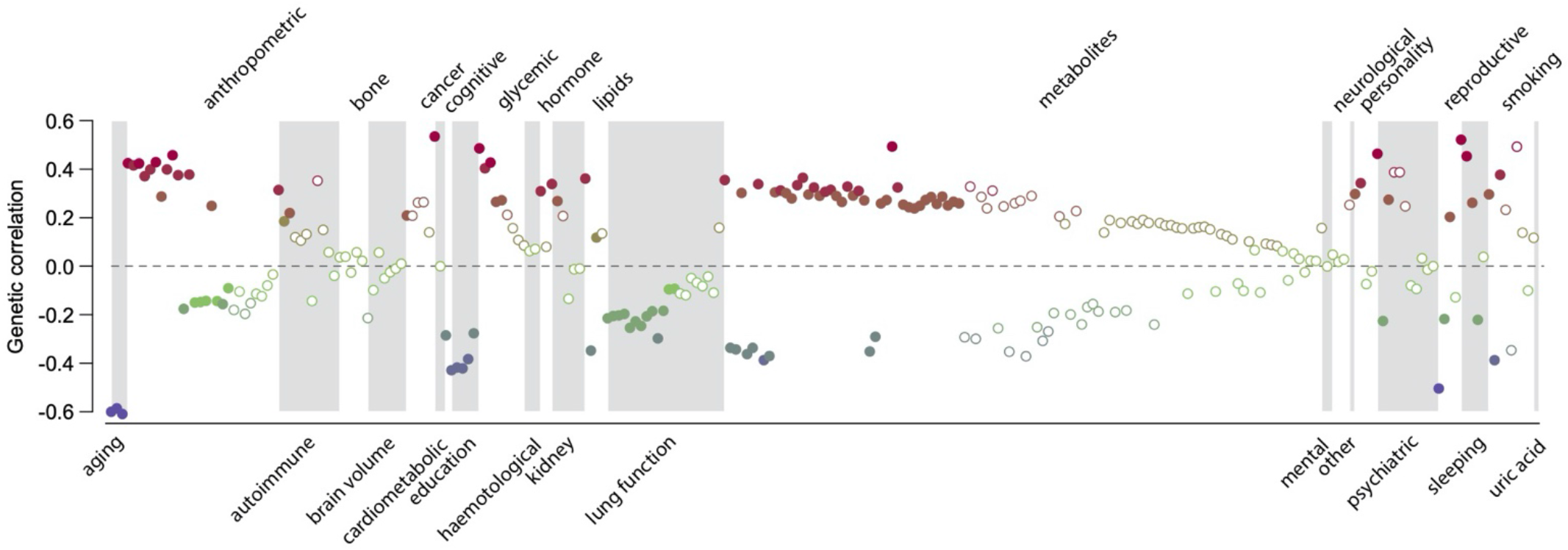
Estimated genetic correlations between medication-use and 257 traits and diseases. Traits displaying significant genetic correlations are displayed as filled symbols. Details can be found in Supplementary Table S3.

**Figure 5.**
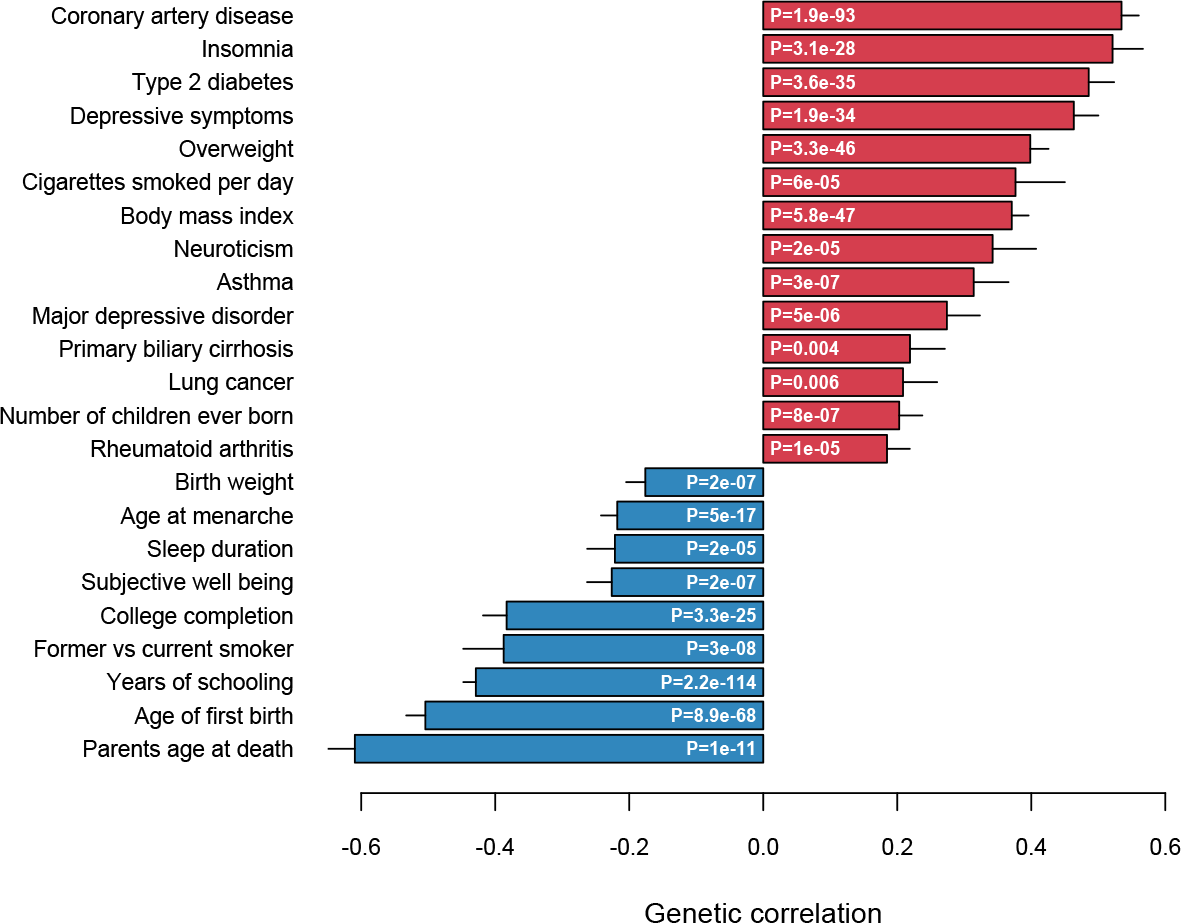
Genetic correlations between medication-use and selected top categories.

## Discussion

In this study, we used data from the UK Biobank to perform a genetic analysis of current medication-use, defined as the total number of different prescription and over-the-counter drugs UKBB participants were taking at the time of the initial assessment. Our aim was to investigate the genetic basis of self-reported medication-use. We identified 59 linkage disequilibrium independent SNPs (*P*< 5 ·10^−8^) associated with current medication-use (Fig. 1). The strongest signal was located within the major histocompatibility complex (MHC); *HLA-DQA1* (lead SNP rs35248896, *P* value = 1.52 ·10^− 46^), which belongs to the MHC class II gene. The MHC region is a large genomic region on chromosome 6 that is associated with more diseases than any other region of the genome (20,25). Additional three loci within MCH reached LD-independently genome-wide significance (Supplementary Table S3), however, the complexity and extreme variant polymorphism, combined with strong LD, within MHC complicates interpretation and disentanglement of individual loci (26). Given the biological involvement of MHC in immune response, it was unsurprisingly that exactly this region was the strongest associated loci.

The associated loci have previously been linked to a large number of different quantitative traits and complex diseases. Since the genetic correlations express the extent to which two measurements reflect what is genetically the same character (27), it is not too surprisingly that we see a good correspondence between the identified loci (Fig. 1) and their previous associations and the significant genetic correlations (Fig. 5, Supplementary Table S4). For example, among our candidate genes (Supplementary Table S3) we have susceptibility loci for diabetes (*PTPN22, CEP68, RREB1, TCF7L2* (28–30)), coronary artery disease (*PSRC1, UNC5C*, *LPLA* (31)), depression (*MAD1L1, YLPM1* (32,33)), and insomnia (*NMT1* (34)). But also for non-disease traits including BMI (*RABGAP1L*, HEYL (35,36)), smoking (NLGN1 (37)), and age at menarche (RBM6 (38)).

The polygenic nature of medication-use was, aside from the large number of identified quantitative trait loci, further supported by the linear association between proportion of genetic variance captured by each autosomal chromosome and the proportion of genetic variants (Fig. 2A), which is similar to what is seen for other complex, polygenic traits (39–43). We observed that low frequent alleles capture more genetic variance than common variants (Fig. 2B), which is similar to what is seen in for example type 2 diabetes (44) and coronary artery disease (42,45). Neurodevelopmental and -degenerative disorders, like schizophrenia, Tourette syndrome and Alzheimer disease, do, however, show the opposite pattern (39,40,46). Therefore, it is not surprisingly that our findings correspond with what is seen for common diseases since the disease prevalence of common diseases in the UKBB follows the population prevalence, and further that the prevalence of mental disorders is too low compared to the population frequency.

Our statistical genetic analysis was performed across any medical condition the included UKBB participants may be diagnosed with as our interest was to study the genetic contributions to variation in medication-use. Thus, our findings could be biased towards the common diseases with the highest disease prevalence, for example through partially shared genetic aetiology. However, this would inevitable imply that some disease groups require numerically more drugs for treatment than other disease groups. Our results clearly demonstrated that individuals with high genetic score for medication-use had an increased medication usage (Fig. 3B), and that this was associated with higher probability of have more disease diagnoses (Supplementary Fig. S5) and having experienced an adverse drug reaction (Supplementary Fig. S7). Thus, it appears, that the individuals with large medication-use has a genetic predisposition towards increased medication demand likely because of poor health from being predisposed to a larger number of different diseases, leading to increased risk of responding negatively to the prescribed medical treatment. Supportive for this, is that we see medication-use to be genetically correlated with known indicators of poor health (Fig. 5). We found, for example, overweight and high body mass index, known to be under strong genetic influence (47), to be positively correlated with medication-use, and these are strongly associated with poor health (48–50). Moreover, the behavioural characteristics smoking and sleep patterns, are also known factors for bad state of health (51,52), which also displayed significant genetic correlations with medication-use (Fig. 5).

This study has a number of limitations. First, despite the information on medication-use was obtained by trained nurses during interviews, the same drug may have been reported under different names, which may limit the accuracy of the analysis. Second, our definition of drug usage also includes vitamins. However, for many diseases, dietary supplements are used in the pharmacological intervention, thus, to prune out vitamins might not more accurately capture the individual’s medication profile. Third, the lack of information on medication duration, dosage and response entails that true pharmacogenomic analysis cannot be performed. Fourth, the self-reported nature of the phenotype may limit the precision; however, studies have shown good concordance between self-reported and electronic health records, both for diseases (53) and for medication-use (54). Fifth, the findings presented here are specific to the UK Biobank participants, which is not representative of the general UK population (55,56), and may not translate to other populations and other health systems.

In conclusion, we have shown that the genetic basis of current medication-use in the UK Biobank among 335,744 individuals appears genetically heterogeneous. We identified 59 independent quantitative trait loci for medication-use and found that 18% of the observed variation could be ascribed to common genetic variants. The genetic heterogenous nature of medication-use was further supported as the genetic variance was spread across the genome, and that the highest accuracy of prediction was observed when including 1.5 million genetic markers. Our findings that medication-use was strongly correlated with smoking behaviour, parents age of death, educational level and insomnia, suggest that the genetic architecture is not biased towards common diseases. Understanding the genetic aetiology of complex diseases has been suggested as a route for improving medical treatment (57,58). Medication-use – as defined in the current study – is an easy quantifiable trait, and because of its genetically corelated nature with many complex traits and diseases, incorporating such information into new multicomponent genetic risk scores (59) could further increase the accuracy of predicting disease predispositions and disease trajectories. From genetic data we can identify those individuals with high medication-use, which concurrently are the most diseased individuals, and they are at an increased risk for adverse drug reaction. Thus, individual medication profiles are likely to be yet another puzzle piece in understanding complex for human diseases, and for providing better medical treatment for future generation.

## Material and Methods

### Genotype and phenotype data

Genetic and phenotypic data were obtained from the United Kingdom Biobank (UKB) (13,60). Data has been collected for more than 500.000 individuals aged 37-73 years. Genotyping details has been described previously (13). To obtain a genetic homogeneous study population we restricted our analyses to unrelated Caucasians, and excluded individuals with more than 5,000 missing genotypes or individuals with autosomal aneuploidy (*n*=335.744). We first converted the imputed genotype probabilities to hard-call genotypes using PLINK2 (--hard-call 0.1) (61), and variants with MAF < 0.01, missing genotype rate >0.05, Hardy-Weinberg equilibrium test *P* value < 1×10^−6^, or imputation info score <0.3 were excluded (*m* = 9.804.629).

We defined current medication-use as the number of different prescription and over-the-counter medicines (Data Field: 20003) the participants were taking regularly at the time of the verbal interview. Any short-term medications, like antibiotics or analgesics were not registered at the interview. From Hohl *et al*. (2013)(19) we obtained a list of ICD10 codes commonly used to describe adverse drug reactions (Supplementary Table S1).

### Genome-wide association study (GWAS) of medication-use

Using PLINK2 (61) we performed a GWAS on medication-use for the entire White-British cohort with sex, age, UKB assessment centre, and the first ten genetic principal components as covariates. To identify high confidence independent associated loci, we performed LD-based clumping in window size of 1000 kb with r^2^<0.01, and for the major histocompatibility complex region we only allowed one significant locus. Lead SNPs within each independent genome-wide significant locus was annotated to nearest gene (genome build GRCh37, hg19) within 2000 kb using Variant Effect Predictor (62).

### Estimation of heritability and genetic correlations

We estimated the proportion of variation in medication-use explained by SNPs 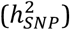 using SumHer (23). We excluded variants within the HLA region (as suggested by the authors of SumHer), and the estimation was performed assuming the LDAK heritability model. In addition, we estimated enrichment for 24 functional categories (obtained from Finucane et al. (2015)(22)), and further partitioned 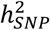 to autosomal chromosomes and minor allele frequency bins.

As it has been shown that SumHer and LD Score regression (63) has similar accuracy in estimation of genetic correlations (23), we used LD Hub to estimate the genetic correlations between medication-use (excluding the HLA region) and 257 quantitative and disease traits (24). To account for multiple testing, we adjusted all *P*-values using Bonferroni (*P*_*adj*_<0.01). Using LD Score regression (63) we also computed the genetic correlation between our definition of medication-use and the previously published genetic analysis of categories of medication traits (16). The univariate LD scores were computed using the 1000 Genomes European data.

### Constructing genetic scores for medication-use

To construct within-cohort genetic scores we reran the GWAS in PLINK2 on five subdivision of the White-British cohort (i.e., a five-fold cross validation scheme). For each set of summary statistics we performed LD clumping (for *r*^2^ < {0.1,0.3,0.5, 0.7, 0.9}) implemented in the R package qgg (41) using a range of *P* value thresholds (0.001, 0.01, 0.05, 0.1, 0.2, 0.3, 0.4, 0.5, 0.7, 0.9, 0.999), and computed the genetic scores as 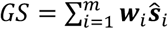, where ***w***_*i*_ is the *i*-th genotype (allelic counts), ***Ŝ***_*i*_ is the estimated SNP effect, and *m* is the number of SNPs left after LD pruning and *P*-value thresholding.

The accuracy of the genetic scores were obtained as the correlation between the number of medications taken of the individuals in the training set and the predicted genetic score. The genetic scores were divided into percentiles, and regression coefficients (*β*) were estimated by linear regression of the number of medications taken on the genetic score percentile relative to the 50^th^ genetic score percentile, adjusted for sex, age, UKB assessment centre, and the first ten genetic principal components.

## Data Availability

The genetic and phenotypic data were obtained from the UK Biobank Resource (ID 31269). Researchers can apply for access through: https://www.ukbiobank.ac.uk/register-apply/

## Funding

This work was supported by a Lundbeck Foundation grant to PDR (R287-2018-735).

### Abbreviations

GWAS: genome-wide association study
ICD10: International Classification of Diseases, version10
SNP: single nucleotide polymorphisms
UKBB: United Kingdom Biobank

## Acknowledgements

All of the computing for this project was performed on the GenomeDK cluster. We would like to thank GenomeDK and Aarhus University for providing computational resources and support that contributed to these research results. The genetic and phenotypic data were obtained from the UK Biobank Resource (ID 31269).

