## Supplementary Material for "The genetic architecture of medication-use"

---

The supplementary material contains the following:

**Supplementary Figures**

| | |
|---|---|
| Figure S1 | Distribution of number of medications used. |
| Figure S2 | Prediction accuracy of medication-use. |
| Figure S3 | Number of SNPs used in constructing genetic scores. |
| Figure S4 | Regression coefficient by percentiles of genetic scores. |
| Figure S5 | ICD10 diagnoses stratified by genetic scores. |
| Figure S6 | Number of individuals within top 50 ICD10 diagnoses. |
| Figure S7 | Medication-use within individuals with adverse drug reactions. |
| Figure S8 | Estimated genetic correlations to other medication traits. |

**Supplementary Tables**

| | |
|---|---|
| Table S1 | List of ICD10 codes for adverse drug reactions. [EXCEL] |
| Table S2 | Number of users per drug. [EXCEL] |
| Table S3 | Genome-wide significant loci for medication-use. [EXCEL] |
| Table S4 | Estimated genetic correlations. [EXCEL] |
| Table S5 | Medication-use among individuals with adverse drug reactions. |
| Table S6 | Estimated genetic correlations to other medication traits. |

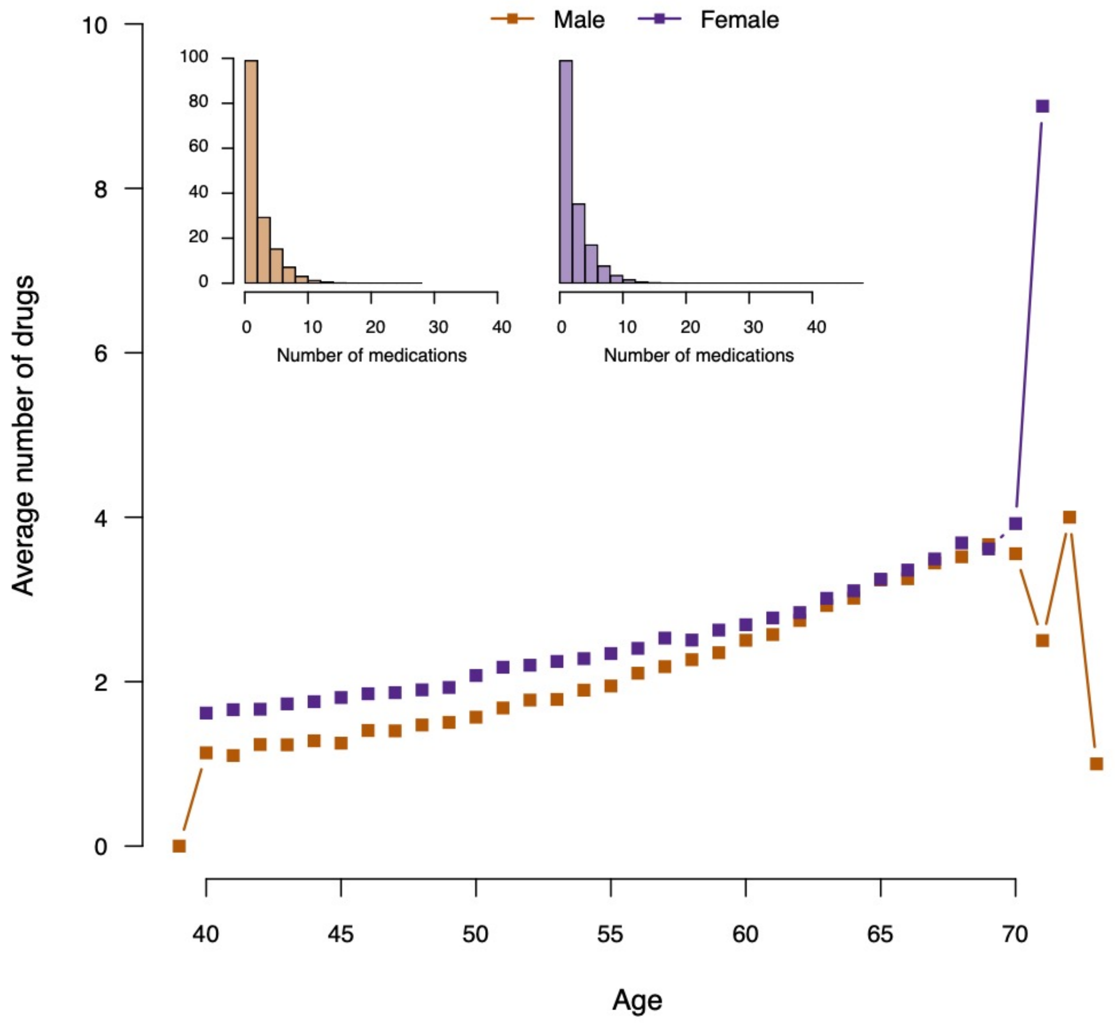


**Figure S1 |** Distribution of number of different drugs each individual in the UKBB White British cohort stratified by age and sex.

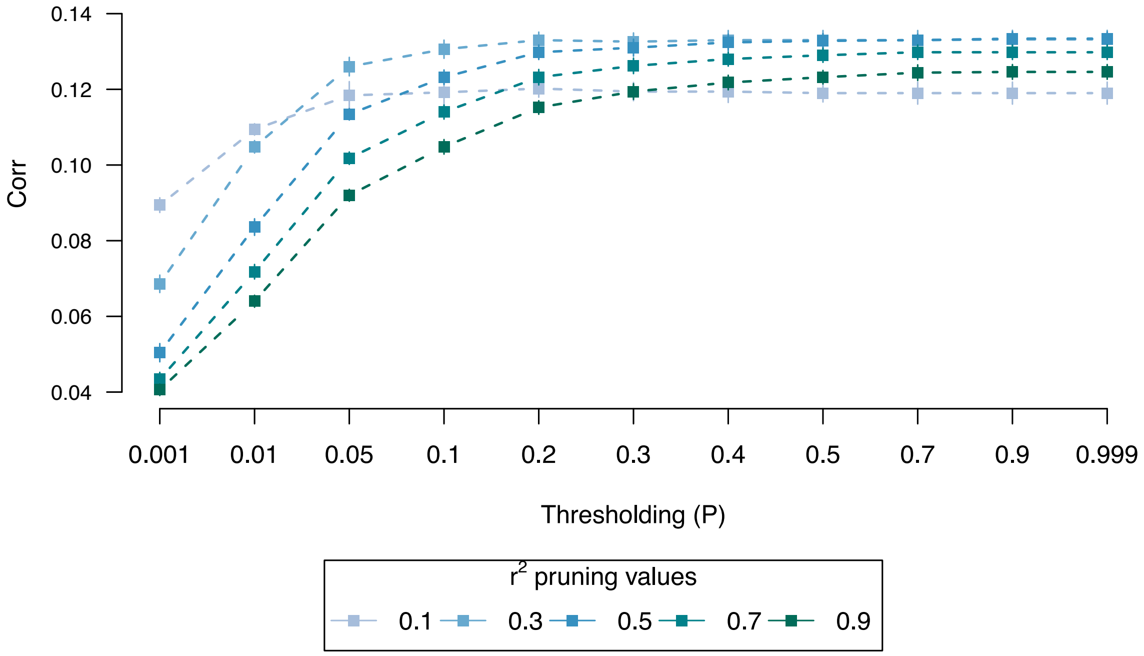


**Figure S2 |** Prediction accuracy (correlation between observed number of medications and the genetic score in the validation data) for different pruning parameters.

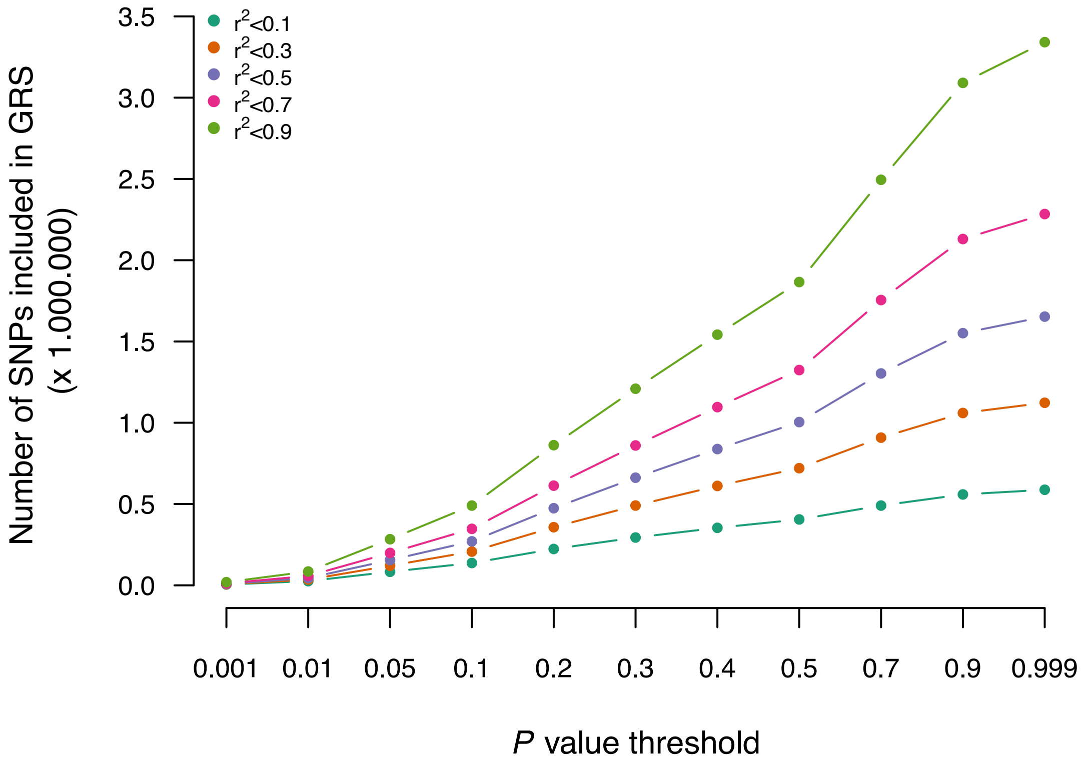


**Figure S3 |** Number of SNPs left after the different parameters for thresholding and LD pruning.

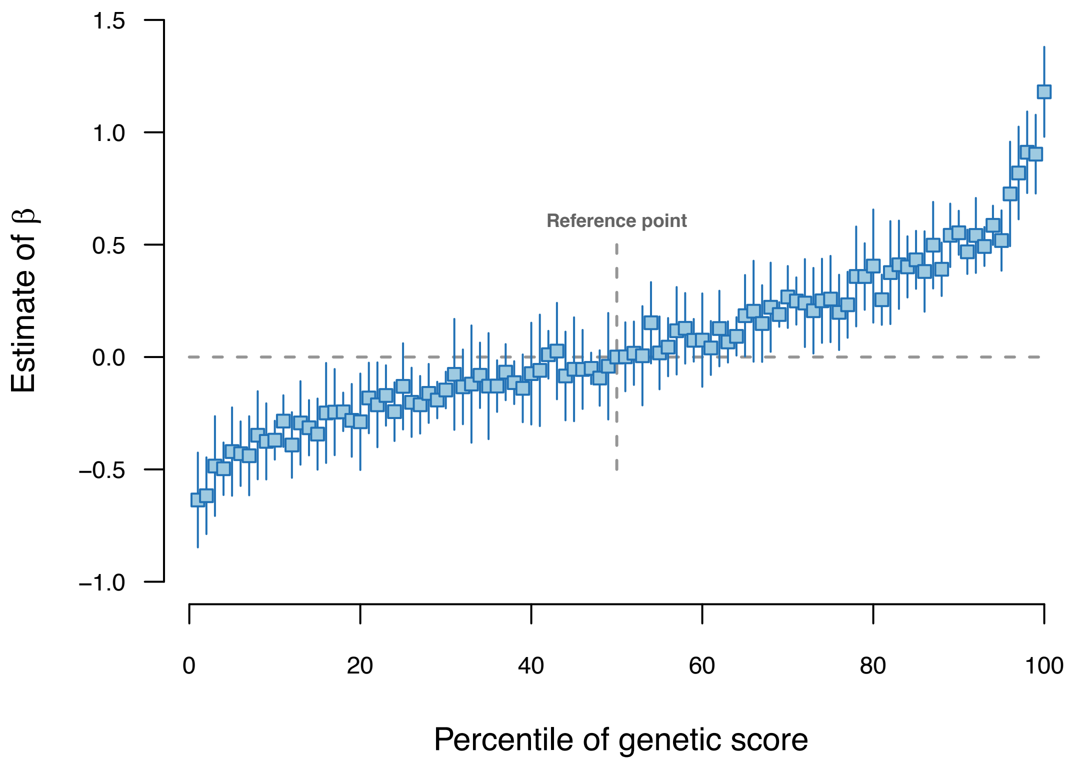


**Figure S4** | Regression coefficient ($\beta$) by percentiles of genetic scores. The $\beta$-estimates were obtained by regressing the number of medications on the genetic score percentile relative to the 50$^{th}$ percentile adjusted for covariates. Results shown are for $r^2<0.5$ and is the mean (and 95% confidence interval) across the five training sets.

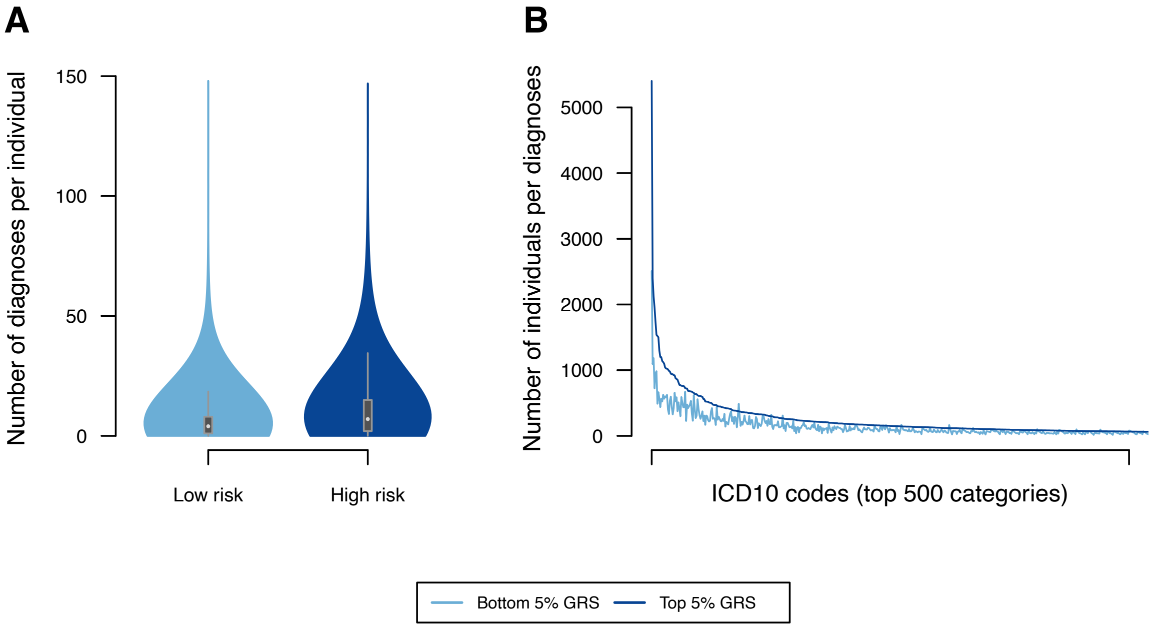


**Figure S5 | A**: Number of ICD10 diagnoses per individual within the bottom 5% (low risk) and in the top 5% (high risk) of genetic scores. **B**: Number of individuals within each ICD10 diagnoses stratified by low and high-risk genetic score. Only the 500 ICD10 codes with most cases are shown.

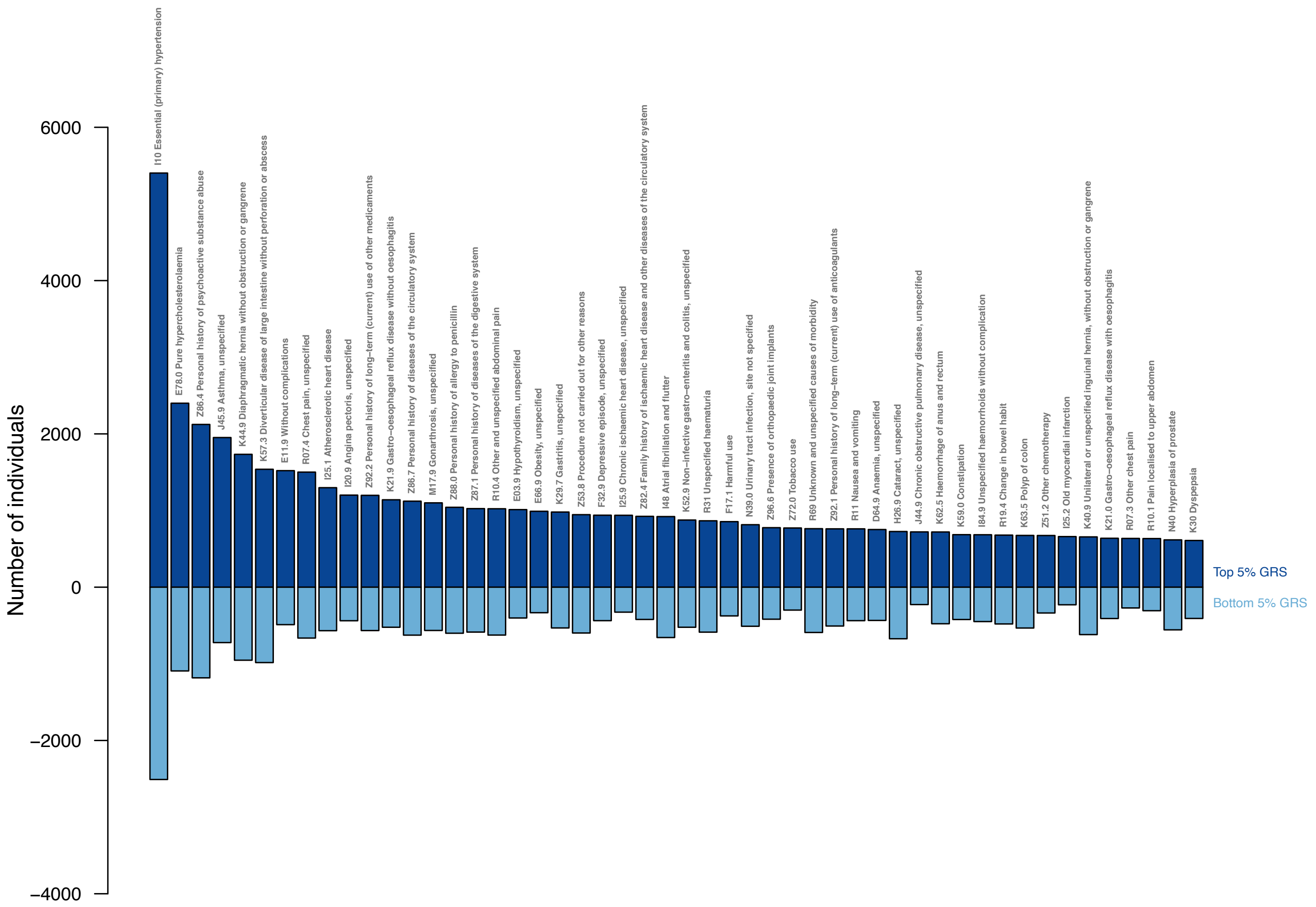


**Figure S6 |** Number of individuals within the top 50 ICD10 diagnoses codes of individuals with the top 5% highest genetic score and bottom 5% lowest genetic score.

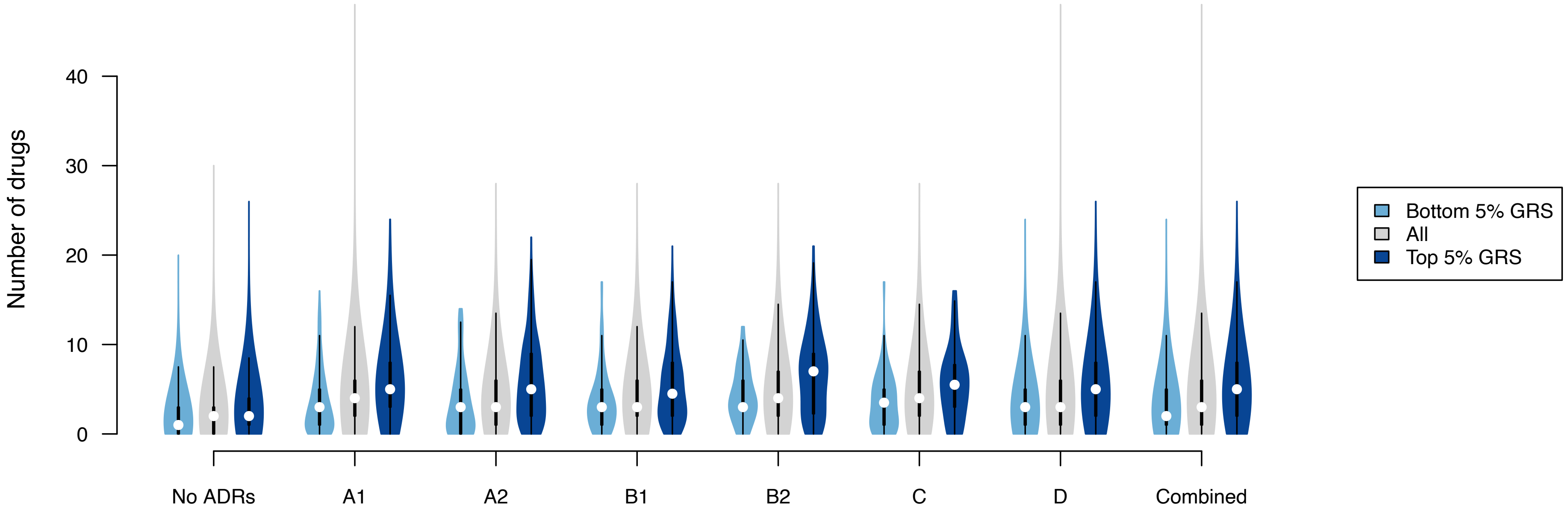


**Figure S7 |** Comparison of number of medications taken by individuals without an experience of any adverse drug reactions (ADR) and individuals suffering from at least one SDR stratified by low and high genetic score. The classification of ADRs are based on Hohl et al (2014; doi:10.1136/amiajnl-2013-002116): A1: The ICD-10 code description includes the phrase 'induced by medication/drug'; A2: The ICD-10 code description includes the phrase 'induced by medication or other causes'; B1: The ICD-10 code description includes the phrase 'poisoning by medication; B2: The ICD-10 code description includes the phrase 'poisoning by or harmful use of medication or other causes'; D: Adverse drug event deemed to be very likely although the ICD-10 code description does not refer to a drug, and D:  Adverse drug event deemed to be likely although the ICD-10 code description does not refer to a drug.

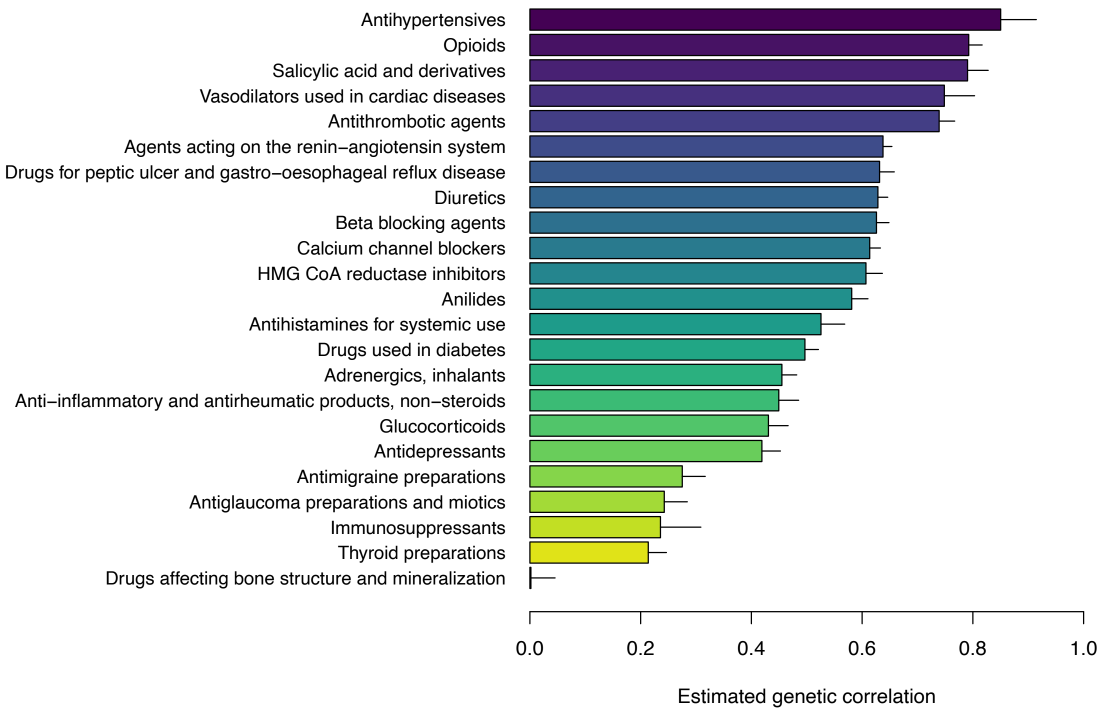


**Figure S8 |** Estimated genetic correlations between medication-use and the 23 categories of medications from Wu et al (2019, doi: 10.1038/s41467-019-09572-5). All estimates are significant, except 'Drugs affecting bone structure and mineralization'. See Supplementary Table S6 for details.

**Table S5 |** Comparison of number of drugs taken for individuals with or without adverse drug reactions (ADRs) for individuals with low or high genetic score for medication-use.

| | Low Risk | | | High Risk | | | |
|---|---|---|---|---|---|---|---|
| **ADR category** | **Mean** | **SD** | **n** | **Mean** | **SD** | **n** | ***P* value*** |
| No ADRs | 1.69 | 2.03 | 15107 | 2.96 | 2.88 | 13988 | $< 2.2 \cdot 10^{-16}$ |
| A1 | 3.43 | 3.37 | 325 | 5.83 | 4.28 | 527 | $< 2.2 \cdot 10^{-16}$ |
| A2 | 3.40 | 3.66 | 94 | 5.81 | 4.51 | 160 | $1.6 \cdot 10^{-6}$ |
| B1 | 3.64 | 3.34 | 107 | 5.45 | 4.17 | 242 | $6.4 \cdot 10^{-6}$ |
| B2 | 4.20 | 2.94 | 29 | 6.60 | 4.53 | 78 | 0.006 |
| C | 3.55 | 3.19 | 58 | 5.57 | 3.96 | 74 | 0.002 |
| D | 3.27 | 3.10 | 1371 | 5.34 | 4.12 | 2288 | $< 2.2 \cdot 10^{-16}$ |
| Combined ADRs† | 3.24 | 3.11 | 1650 | 5.28 | 4.08 | 2774 | $< 2.2 \cdot 10^{-16}$ |
| Multiple ADRs‡ | 3.81 | 3.52 | 274 | 6.19 | 4.48 | 478 | $5.6 \cdot 10^{-16}$ |

* adjusted for age and sex

† Having at least one ADR within one of the six ADR categories

‡ Having more than one ADR within one of the six ADR categories

**Table S6 |** Estimated genetic correlations between medication-use and the 23 categories of medications from Wu et al (2019, doi: 10.1038/s41467-019-09572-5).

| **Code** | **Name** | **Genetic correlation** | **Standard error** | ***P* value** |
|---|---|---|---|---|
| A02B | Drugs for peptic ulcer and gastro-oesophageal reflux disease | 0.63 | 0.027 | 5.26E-125 |
| A10 | Drugs used in diabetes | 0.45 | 0.024 | 4.09E-92 |
| B01A | Antithrombotic agents | 0.74 | 0.028 | 2.14E-152 |
| C01D | Vasodilators used in cardiac diseases | 0.75 | 0.055 | 9.17E-43 |
| C02 | Antihypertensives | 0.85 | 0.064 | 9.59E-40 |
| C03 | Diuretics | 0.63 | 0.018 | 7.90E-274 |
| C07 | Beta blocking agents | 0.63 | 0.023 | 1.83E-168 |
| C08 | Calcium channel blockers | 0.61 | 0.019 | 3.43E-219 |
| C09 | Agents acting on the renin-angiotensin system | 0.64 | 0.016 | 0.00E+00 |
| C10AA | HMG CoA reductase inhibitors | 0.61 | 0.030 | 1.03E-92 |
| H03A | Thyroid preparations | 0.21 | 0.033 | 6.89E-11 |
| L04 | Immunosuppressants | 0.24 | 0.073 | 0.0012 |
| M01A | Anti-inflammatory and antirheumatic products, non-steroids | 0.45 | 0.036 | 5.57E-36 |
| M05B | Drugs affecting bone structure and mineralization | 0.002 | 0.044 | 0.9674 |
| N02A | Opioids | 0.79 | 0.024 | 8.85E-233 |
| N02BA | Salicylic acid and derivatives | 0.79 | 0.037 | 6.34E-100 |
| N02BE | Anilides | 0.58 | 0.029 | 2.98E-87 |
| N02C | Antimigraine preparations | 0.28 | 0.042 | 3.51E-11 |
| N06A | Antidepressants | 0.42 | 0.034 | 6.28E-36 |
| R03A | Adrenergics, inhalants | 0.46 | 0.027 | 1.16E-64 |
| R03BA | Glucocorticoids | 0.43 | 0.036 | 1.05E-33 |
| R06A | Antihistamines for systemic use | 0.53 | 0.043 | 1.74E-34 |
| S01E | Antiglaucoma preparations and miotics | 0.24 | 0.042 | 5.22E-09 |
